# Identification of Subgroups of Individuals Experiencing Patellofemoral Pain with Kinematic and Kinetic Features During Overground Running

**DOI:** 10.64898/2026.01.13.26344049

**Authors:** Ross J. Brancati, Wouter Hoogkamer, Douglas N. Martini, Katherine A. Boyer

## Abstract

Patellofemoral pain (PFP) is a common running related injury associated with several biomechanical factors such as altered kinematics and kinetics across lower extremity joints. Prior research suggests mechanisms for PFP may differ within those affected, indicating that subgroups characterized by unique factors might exist. Subgroups have previously been found using unsupervised machine learning (ML) models based on clinical tests, imaging, and/or biomechanical stair negotiation variables, however clusters have not yet been identified with 3-dimensional running mechanics. Therefore, the purpose of this study was to leverage an unsupervised ML algorithm to identify clusters of individuals experiencing unique PFP related biomechanical factors and furthermore characterize features of each subgroup. 40 participants experiencing PFP and 20 healthy controls completed overground running trials in a motion capture environment. Kinematic and kinetic variables were reduced with principal component (PC) analysis. PC-based features were fed into a k-means clustering model optimized on a measure of cluster purity. Four unique subgroups were identified, three of which contained mostly individuals experiencing PFP. Each subgroup displayed distinct kinematic and/or kinetic features of running gait related to development of PFP. The first PFP subgroup was characterized by excessive knee abduction and rotation moments, and reduced rearfoot inversion moments. The second PFP subgroup displayed greater knee joint frontal plane loading rates and rearfoot eversion angles. The third PFP subgroup was characterized by greater hip adduction, hip rotation, and knee abduction angles. Identifying and characterizing subgroups is a key step for developing targeted treatments that may be more effective for mitigating PFP.

**Highlights:**

- Three distinct subgroups were identified with a clustering model and principal component-based features, each with unique kinematic and kinetic patterns during overground running.
- Key features for differentiating subgroups included the knee abduction moment, rearfoot inversion moment, knee frontal plane loading rate, rearfoot eversion angle, hip adduction angle, hip rotation angle, and knee abduction angle.
- Each subgroup may benefit from targeted intervention strategies as opposed to generalized treatment procedures.

## Introduction

Patellofemoral pain (PFP) syndrome has an estimated prevalence of 23% in the adult population and is characterized by pain beneath or surrounding the patella (1–4). PFP symptoms are typically exacerbated by activities that repetitively load the patellofemoral joint, such as running (1). Thus, PFP can lead to decreased participation in physical activity, diminished quality of life, and may also increase risk of development of patellofemoral osteoarthritis (5–8). Treatments for PFP are not always effective, as demonstrated by 21-40% of patients reporting poor outcomes after intervention (9,10). Additionally, a prior study showed that over 50% of patients experienced symptoms up to eight years after initial treatment (11). Poor outcomes associated with current generalized interventions suggest that targeted interventions are necessary (12,13).

The etiology of PFP is complex. There are anatomical, biomechanical, psychological, behavioral, and social factors that contribute to pain development (14). Identifying and understanding such factors has been a key focus of research to support development of more effective treatment plans (13,15). Diminished joint contact area and increased joint reaction forces, the two primary mechanisms of PFP, are related to kinematic and kinetic adaptations local, distal, or proximal to the knee joint (14). For example, prior cross-sectional studies have found that those with PFP exhibited greater hip adduction and internal rotation angles, knee abduction angles, knee abduction impulse, and rearfoot eversion during stance phases of walking and running compared to healthy controls (16–20). However, a recent meta-analysis only found moderate evidence for reduced peak knee flexion angles and limited evidence of reduced peak knee extension moments in those with PFP compared to healthy controls, and the standardized mean differences for both biomechanical variables were small (21), indicating conflicting prior findings from cross-sectional studies. A potential explanation of the lack of consistency across studies is the existence of PFP subgroups with unique movement adaptations leading to altered joint mechanics. Evidence for distinct subgroups among individuals with PFP emerged from an initial study that observed three subgroups characterized by *knee valgus*, *excessive hip abduction*, and *normal gait patterns* during treadmill running (17).

While this initial study (17) identified these PFP subgroups post-hoc as a way to understand high variability in the study outcomes, more recent work has used unsupervised machine learning (e.g., clustering) approaches to uncover subgroups. One of the first data driven studies leveraged hierarchical clustering and latent profile analysis with data from seven clinical tests, demonstrating potential use in clinical settings (22). This study found three distinct subgroups, described as 1) weak and tighter, 2) weak and overpronated, and 3) strong (22). Drew et al. (2019) used flexibility measures, imaging techniques, strength assessments, and foot posture metrics to distinguish four subgroups defined as 1) strong, 2) pronated and malaligned, 3) weak, and 4) active and flexible. Few studies have focused on dynamic measures, however, one study used pelvic acceleration profiles from treadmill running in a hierarchical clustering analysis, showcasing the potential of wearable sensor-based clustering (24). In that study, two distinct female subgroups, but no distinct male subgroups were identified. Post-hoc analysis revealed that subgroups displayed unique kinematic characteristics at the foot, knee, and hip. While this study was advantageous in that findings could be easily translated to wearable sensors, the complexity of biomechanical factors that influence patellofemoral joint stress were not considered in the clustering approach. While prior research demonstrates the existence of subgroups, no studies have identified them with kinematic and kinetic variables known to contribute to PFP during pain exacerbating activities such as running using data driven approaches.

To date there has been limited success using unsupervised clustering to identify biomechanical subgroups associated with injury status. One study applied hierarchical clustering to discrete kinetic variables associated with running injury and identified two subgroups; however, injured and non-injured runners were similarly distributed across clusters, potentially due to heterogeneity in injury pathology or limitations of the clustering approach (25). Similarly, another study found that kinematic features did not effectively uncover any association between subgroups and injury status or location (26). Although both studies used hierarchical clustering to identify biomechanically distinct subgroups, neither approach explicitly incorporated injury status or attempted to constrain the formation of a homogeneous healthy control subgroup. As a result, potential injury-specific gait patterns may have been obscured by both heterogeneous injury presentations and purely unsupervised clustering methodologies. Therefore, specific injury pathology and methodological considerations for identifying clusters should be accounted for in future studies attempting to identify PFP subgroups.

There is now support for the existence of subgroups in PFP, but the best approach to identify these subgroups and their defining characteristics remain unclear. Running injury subgrouping may have important implications for treatment of individuals experiencing PFP. Prior work has shown that a posthoc analysis of baseline running kinematics and self-reported measures can differentiate treatment responders from non-responders (10,27). Furthermore, targeting treatments at defined subgroups can result in superior outcomes to generalized approaches (22). Together, these findings highlight the need to better characterize biomechanical PFP subgroups to enable more effective, individualized intervention strategies.

The purpose of this study was to identify subgroups within the PFP population using an unsupervised machine learning approach with kinematic and kinetic features and furthermore characterize unique biomechanics within each subgroup while accounting for healthy, uninjured movement patterns. It was hypothesized that up to four distinct subgroups would exist, each with unique biomechanical patterns related to development of PFP. The number of clusters is based on prior subgrouping studies; however, the characteristics of each cluster cannot be speculated due to a lack of prior research in this area. Identifying and characterizing PFP subgroups could provide important information needed to develop more targeted, personalized treatment approaches in this population.

## Materials and Methods

### Participants

20 healthy, pain free individuals and 40 individuals symptomatic for PFP were recruited for this study. All participants were physically active, between the ages of 18-40, and had no history of traumatic lower extremity injury or neurological pathology. Participants with PFP were required to have a clinical diagnosis by self-report or reported symptoms of PFP but not symptoms of other running injuries (1). Each participant was required to pass the Physical Activity and Readiness Questionnaire (28). All study details were approved by the University of Massachusetts Amherst Institutional Review Board and each participant completed an informed consent before participating in the study.

### Surveys and Instrumentation

Participants completed a survey with 23 questions from the REPORT-PFP Checklist developed by the International Patellofemoral Research Network (29) at the start of the visit. Each participant was also given a standardized shoe (Brooks T7 Racer, Seattle, WA) and completed a warmup consisting of stretching and a five-minute treadmill run at a preferred speed.

Participants were then fit with reflective markers on the left and right anterior superior iliac spine (ASIS) and posterior superior iliac spine (PSIS). Markers were also placed unilaterally on the lateral femoral condyle, medial femoral condyle, lateral tibial plateau, medial tibial plateau, lateral malleolus, medial malleolus, first metatarsal head, second metatarsal head, and fifth metatarsal head. Additionally, plates of four non-colinear markers were placed on the lateral aspect of the thigh one third of the distance from the lateral femoral condyle to the greater trochanter and on the lateral aspect of the shank one third of the distance from the lateral tibial plateau to the lateral malleolus. Markers were also placed on the calcaneus, lateral and inferior to the calcaneus, and medial and inferior to the calcaneus to track foot motion. In the symptomatic PFP group, the testing limb was selected as symptomatic limb. In the case of bilateral pain, the more severely affected limb was tested. For the healthy individuals, the dominant limb was tested.

### Data Collection

Data collection began with a static trial recorded with 11 motion capture cameras (Oqus 300, Qualisys, Gothenburg, Sweden) while the participant stood on a force plate (AMTI, Watertown, MA). Upon completion of the static trial, ten successful overground running trials were completed on a 22 m long runway: five at the participant’s preferred speed and five at a set speed of 3.16 m/s. Preferred running speed was calculated as the average of three overground running trials before starting data collection. A successful trial consisted of a clean foot strike on a force plate where the speed was within 5% of the desired running speed. For each running trial, motion capture marker trajectories and force plate data were recorded at 128Hz and 1920Hz, respectively. Running speed was monitored by timing gates placed three meters before and after the force plate.

### Data Processing

The pelvis segment was modeled as a CODA pelvis using the ASIS and PSIS markers, and hip joint centers were defined with Bell regression equations (30). Proximal and distal thigh and shank anatomical coordinate systems were defined as previously described (31). The foot anatomical coordinate system was located at the midpoint of the lateral and medial malleoli, and the longitudinal axis was defined as a vector from this location to the midpoint of the first and fifth metatarsal heads. The thigh and shank segment were tracked with plates of four non-colinear markers, and the foot segment motion was tracked using the three heel markers and three metatarsal head markers. Segment masses, moments of inertia, and center of mass locations were defined according to Dempster (1955) and Hanavan (1964). Three dimensional joint angles and pelvis orientations were calculated with Cardan XYZ (flexion-adduction-rotation) rotation sequences. Internal joint moments were computed with a Newton-Euler based inverse dynamics approach (34) and normalized as percent of body weight • height (%BW•H). Moments were resolved to distal segments, and positive forces were defined in the superior, lateral, and anterior directions. Kinematic and kinetic, including 3D ground reaction force (GRF), data were filtered with low-pass Butterworth filters with cutoff frequencies of 8 and 50 Hz, respectively (16,35). GRFs were normalized as percent of body weight (%BW). All kinematic and kinetic processing was performed in Visual 3D (C-Motion Inc., Germantown, MD).

### Data Reduction

All variables were normalized to the stance phase of the gait cycle (36), resulting in 51 data points for all variables except GRF signals which were normalized to 101 data points. Our prior work with a supervised classification approach did not reveal features during swing phase that differentiate PFP from healthy runners (37). Independent matrices were constructed for each variable, where each row represented a trial for either the preferred or set running speed, and each column represented a timepoint of the stance phase. All matrices were reduced with principal component analysis (PCA) (37). PCs that explained 95% of the cumulative variance were retained for further analysis (38). Data was projected onto PCs to compute PC scores, resulting in 1-4 retained PCs for each variable (37,39,40). These scores were concatenated to generate a single feature matrix where each column represented a variable-specific PC, and each row represented the respective score for a trial. Participant-wise means of PC scores were calculated for preferred speed running trials. Both the preferred and set running speed trials were used to improve the stability of PCs.

### Clustering

A k-means clustering model with a wrapper feature selection approach, a method that selects features resulting in optimal clusters using a metric of cluster performance (41), was used to identify clusters. This approach is described in detail in the following sections starting with defining the clustering performance evaluation metric: *cluster purity index*.

### Defining Cluster Purity Index and Gini Impurity

Cluster purity index (CPI) was defined to measure clustering performance at each iteration of the wrapper approach (Equation 1). Optimizing clustering performance with CPI facilitated resultant clusters with the largest possible number of healthy individuals in a single cluster, and symptomatic individuals distributed amongst remaining clusters. The first term of CPI has a maximal score of 1 when all healthy individuals are assigned to a pure, single cluster, while the second term applies a penalty when the cluster contains symptomatic individuals. Therefore, by maximizing CPI amongst several feature sets, one cluster would ideally contain all, or most of, the healthy participants and few to no symptomatic participants, while the other clusters would contain mostly symptomatic participants with few to no healthy participants. Healthy participants are key to cluster optimization. Without them, the risk of identifying clusters with characteristics unrelated to PFP increases. For example, clusters identified without a healthy group could be simply characterized by factors unrelated to PFP such as rearfoot, midfoot, or forefoot striking patterns. Optimizing clusters using CPI provides a more objective criterion for cluster selection than approaches that rely on subjective decisions about the number of clusters.

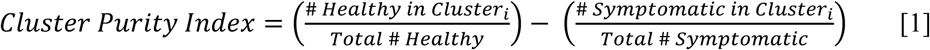

Gini impurity and weighted Gini impurity were also computed for the final cluster assignments (42). The weighted Gini impurity index provides a measure of the overall purity across clusters, where the weighting factor was determined by the number of individuals within each group. Detailed calculations are provided in the supplementary methods.

### Wrapper Method for Feature Selection

Sequential backward selection was used to select features, and clustering performance was evaluated for each feature set with CPI (43). The feature selection approach started with all features and iteratively removed features. CPI was calculated for each feature set and the features that maximized CPI were retained for further analysis. The initial feature set was composed of participant wise PC scores from preferred running speed trials. This feature selection approach, including the algorithm, is described in greater detail in the supplementary methods section.

### Statistical Analysis for Cluster Characterization and Feature Interpretation

One-way analysis of variance (ANOVA) with *α* = 0.05 was used to test for differences between clusters for PC scores of kinematic and kinetic variables and continuous characteristic variables (age, BMI, preferred running speed), with *η*^2^ calculated as a measure of effect size. Post-hoc testing was conducted with Tukey’s honestly significant difference tests and Cohen’s d (*d*) effect sizes were calculated where appropriate. Chi-square tests of independence with *α* = 0.05 were used to test for differences in categorical characteristics and clinical variables in the REPORT-PFP Checklist between clusters, with post-hoc pairwise comparisons performed where appropriate. All statistical assumptions were confirmed. PCs were interpreted with a graphical-visual method where discriminant vectors are compared for 5^th^ and 95^th^ percentile PC scores, which is described in detail elsewhere (37,44). Data processing and statistical analysis were performed in python 3.9 (Python Software Foundation, Wilmington, DE) using appropriate libraries.

## Results

### Clustering Results and Group Characteristics

Clustering analysis revealed four distinct subgroups within our sample with a CPI of 0.475 and a weighted Gini impurity of 0.330, indicating moderately pure clusters. The resulting Dunn index (45) was 0.413, indicating moderately separated and compact clusters. Cluster 1 (C1: 12 PFP, 1 healthy) and 4 (C4: 9 PFP, 1 healthy) contained mostly individuals with PFP. Cluster (C2: 7 PFP, 13 healthy) was the cluster with the majority of healthy individuals. Lastly, cluster (C3: 12 PFP, 5 healthy) had a split of ∼70% PFP and ∼30% healthy. The proportion of females in each cluster was different (*χ*^2^= 8.64, *p* = 0.034, *df* = 3). Post-hoc testing revealed a greater number of females in C4 compared to C3 (*p* = 0.014). BMI was significantly different between clusters (*F* = 6.301, *p* < 0.001, *η*^2^ = 0.250, Table 1), where C4 had a higher BMI than C1 (*p* = 0.017, *d* = 0.977), C2 (*p* < 0.001, *d* = 1.423), and C3 (*p* = 0.042, *d* = 0.845). The preferred running speed was significantly different between clusters (*F* = 5.154, *p* = 0.003, *η*^2^ = 0.216, Table 1), where C4 ran slower than C2 (*p* = 0.001, *d* = 1.48). There were no significant differences between ages of clusters (*F* = 1.05, *p* = 0.377, *η*^2^ = 0.05) or the variables in the REPORT-PFP Checklist (Supplementary Tables 1 and 2).

**Table 1:** Participant characteristics, changes in pain, and Gini impurity for each cluster. Age, BMI, and preferred running speed shown as mean (standard deviation)

| Cluster Label | # Participants in Cluster | # Female | Age (years) | BMI (kg*m<sup>-2</sup>) | Preferred Running Speed (m/s) | Gini Impurity |
| --- | --- | --- | --- | --- | --- | --- |
| C1 | 13 | 7 | 28.7 (7.95) | 23.7 (2.55)* | 3.57 (0.44) | 0.142 |
| C2 | 20 | 11 | 25.4 (6.98) | 22.3 (2.30)* | 3.85 (0.57)* | 0.455 |
| C3 | 17 | 4* | 25.1 (6.86) | 25.1 (6.87)* | 3.55 (0.58) | 0.415 |
| C4 | 10 | 8 | 28.3 (5.92) | 28.2 (6.34) | 3.01 (0.50) | 0.180 |

\* significant difference from C4

**Table 2:** Explained variance, mean and standard deviation (SD) of PC scores, ANOVA results, and post-hoc testing results for the kinematic and kinetic variables that characterized C1.

| Variable | Explained<br>Variance | PC Score Mean (SD) |  |  |  | ANOVA Results |  |  | Post-Hoc Testing Results |  |  |  |  |  |
| --- | --- | --- | --- | --- | --- | --- | --- | --- | --- | --- | --- | --- | --- | --- |
| | | C1 | C2 | C3 | C4 | F | p | $\eta^2$ | C1 vs.<br>C2 | C1 vs.<br>C3 | C1 vs.<br>C4 | C2 vs.<br>C3 | C2 vs.<br>C4 | C3 vs.<br>C4 |
| Knee<br>Abduction<br>Moment<br>PC1 | 85.40% | 0.089<br>(0.092) | -0.022<br>(0.068) | -0.037<br>(0.069) | -0.005<br>(0.098) | 6.698 | 0.001 | 0.264 | p = 0.002<br>d = 1.372 | p < 0.001<br>d = 1.529 | p = 0.043<br>d = 0.945 | p = 0.938<br>d = 0.224 | p = 0.953<br>d = -0.204 | p = 0.756<br>d = -0.384 |
| Knee<br>Rotation<br>Moment<br>PC1 | 97.60% | 0.170<br>(0.056) | -0.049<br>(0.035) | -0.103<br>(0.043) | 0.033<br>(0.11) | 52.502 | < 0.001 | 0.738 | p < 0.001<br>d = 4.745 | p < 0.001<br>d = 5.340 | p = 0.001<br>d = 1.555 | p = 0.054<br>d = 1.336 | p = 0.006<br>d = -1.147 | p < 0.001<br>d = -1.750 |
| Rearfoot<br>Inversion<br>Moment<br>PC1 | 95.11% | 0.132<br>(0.061) | -0.054<br>(0.044) | -0.094<br>(0.052) | 0.040<br>(0.106) | 33.908 | < 0.001 | 0.645 | p < 0.001<br>d = 3.488 | p < 0.001<br>d = 3.885 | p = 0.009<br>d = 1.047 | p = 0.267<br>d = 0.814 | p = 0.003<br>d = -1.285 | p < 0.001<br>d = -1.693 |

### Characterizing Cluster 1 (C1)

There was a significant difference for the first PC of the knee abduction moment, where the PC score of C1 was significantly higher than C2, C3, and C4 (Table 2, Figure 1.A1). indicating a larger moment over the stance phase (Figure 1.A2). There was also a significant difference for the first PC of the knee rotation moment, where the PC score of C1 was significantly higher than C2, C3, and C4 (Table 2, Figure 1.B1), indicating a larger moment over the stance phase (Figure 1.B2). Additionally, there was a significant difference for the first PC score of the rearfoot inversion moment, where the PC score of C1 was significantly higher than C2, C3, and C4 (Table 2, Figure 1.C1), indicating a smaller overall magnitude during stance (Figure 1.C2). Of note, the PC scores of C4 for the knee rotation and rearfoot inversion moments were significantly higher than C2 and C3.

**Figure 1:**
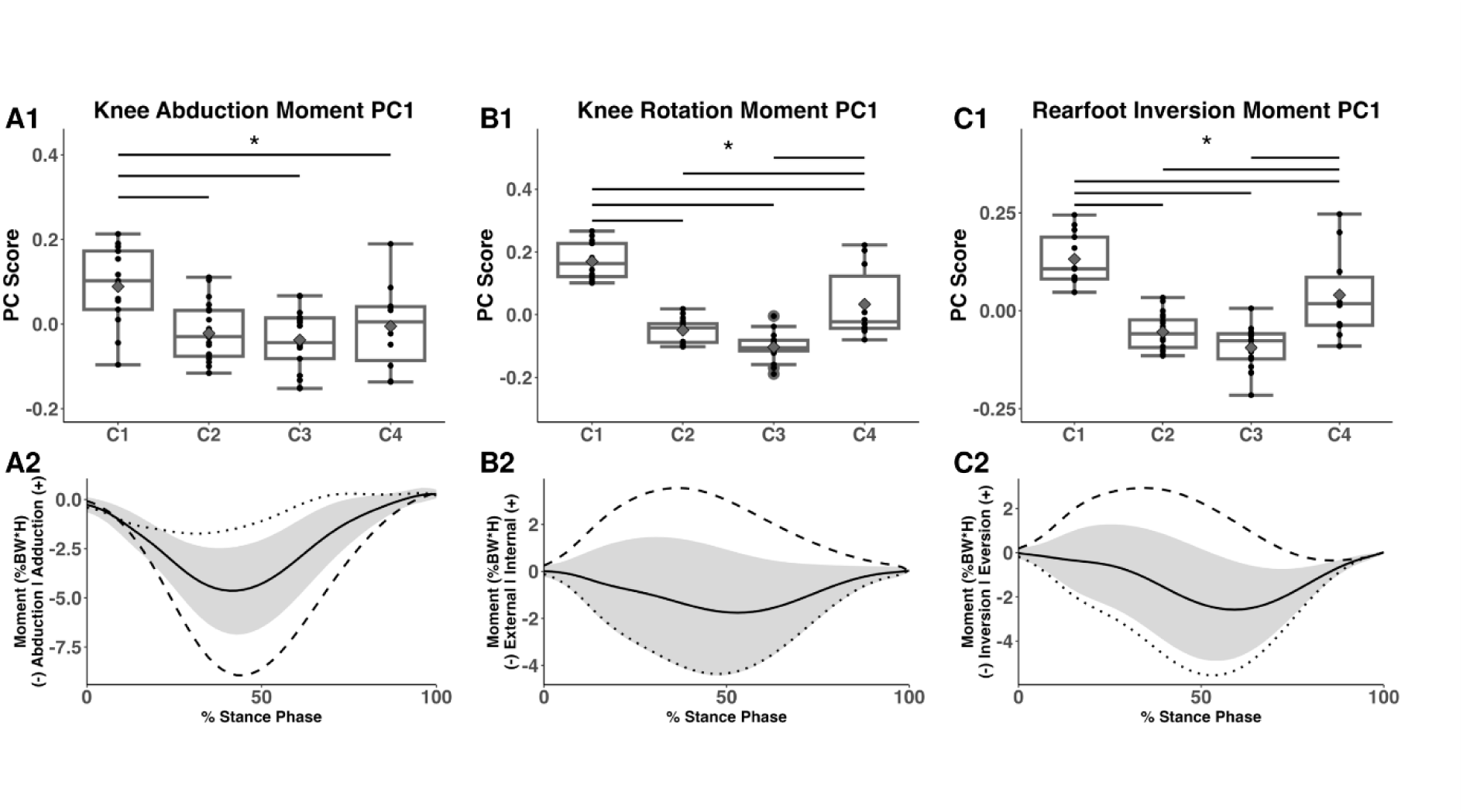
Box plots (top) and interpretation plots (bottom) for the primary kinematic and kinetic variables that characterized C1. Gray diamonds represent group means and black dots represent individual observations. * significant difference from other groups indicated by significance bars. In the interpretation plots, the dotted and dashed lines represent the discriminant vectors for the 5^th^ and 95^th^ percentile scores, respectively. PCs were interpreted by identifying deviations from the mean time-series signal shown in black with standard deviation represented by the gray shaded region.

Additional variables including the first PCs of the rearfoot rotation and hip rotation moments were also unique to C1. For both, C1’s score was significantly higher than C2, C3, and C4 (Supplementary Table 3, Supplementary Figure 2A). C4’s scores were also significantly higher than C2 and C3. Both variables indicated larger moments over the stance phase (Supplementary Figure 2B).

### Characterizing Cluster 3 (C3)

There was a significant difference for the second PC of the knee abduction moment, where the PC score of C3 was significantly lower than C1 and C4, but not C2 (Table 3, Figure 2.A1), indicating an earlier peak moment (Figure 2.A2). Of note, the PC score of C2 was also significantly lower than C1. There was also a significant difference for the first PC of the medial-lateral GRF, where the PC score of C3 was significantly higher than C1 and C4, but not C2 (Table 3, Figure 2.B1), indicating a more medially directed GRF over the stance phase (Figure 2.B2). Additionally, there was a significant difference for the second PC of rearfoot eversion angle, where the PC score of C3 was significantly higher than C1, C2, and C4 (Table 3, Figure 2.C1), indicating larger overall range of motion over the stance phase (Figure 2.C2)

**Figure 2:**
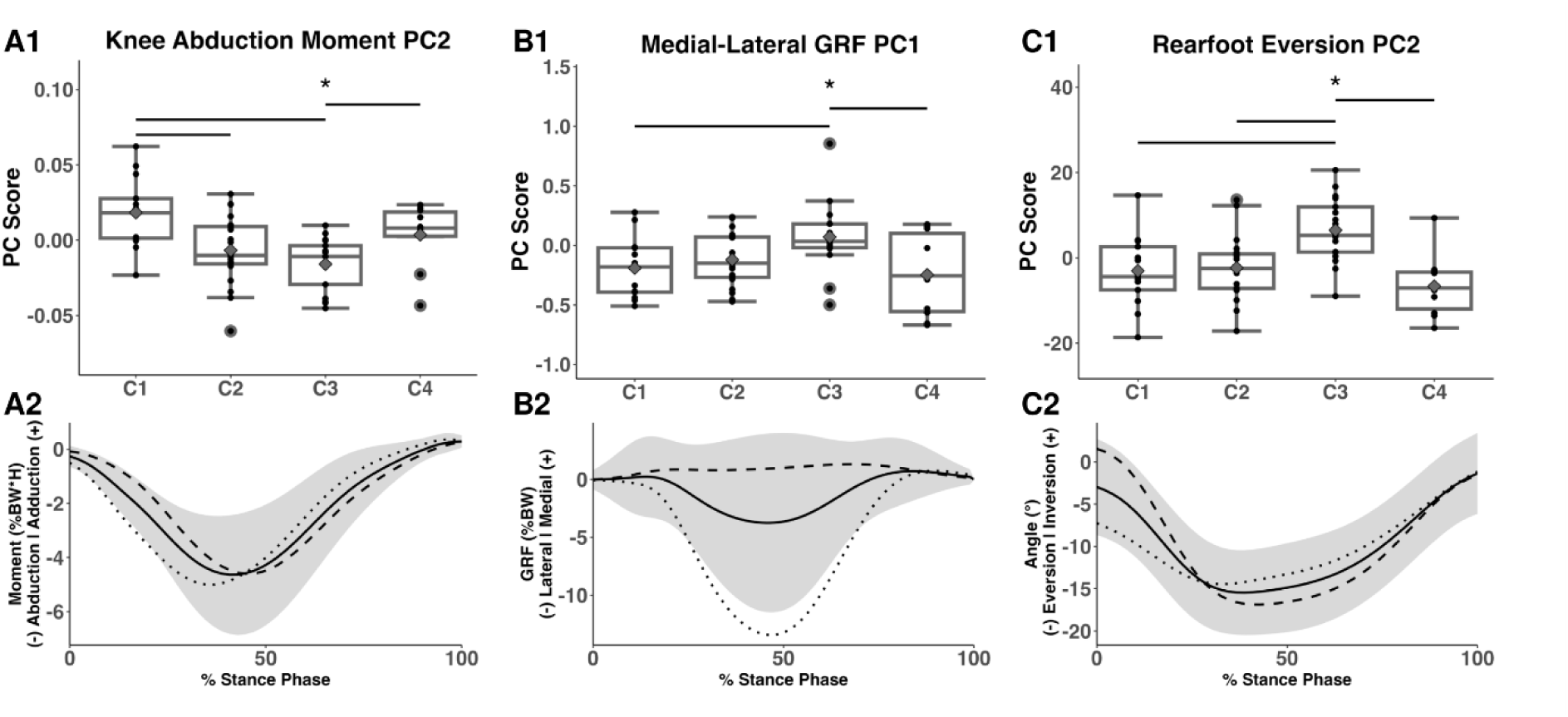
Box plots (top) and interpretation plots (bottom) for the primary kinematic and kinetic variables that characterized C3. Gray diamonds represent group means and black dots represent individual observations. * significant difference from other groups indicated by significance bars. In the interpretation plots, the dotted and dashed lines represent the discriminant vectors for the 5^th^ and 95^th^ percentile scores, respectively. PCs were interpreted by identifying deviations from the mean time-series signal shown in black with standard deviation represented by the gray shaded region.

**Table 3:** Explained variance, mean and standard deviation (SD) of PC scores, ANOVA results, and post-hoc testing results for the kinematic and kinetic variables that characterized C3.

| Variable | Explained Variance | PC Score Mean (SD) |  |  |  | ANOVA Results |  |  | Post-Hoc Testing Results |  |  |  |  |  |
| --- | --- | --- | --- | --- | --- | --- | --- | --- | --- | --- | --- | --- | --- | --- |
| | | C1 | C2 | C3 | C4 | F | p | $\eta^2$ | C1 vs. C2 | C1 vs. C3 | C1 vs. C4 | C2 vs. C3 | C2 vs. C4 | C3 vs. C4 |
| Knee Abduction Moment PC2 | 7.38% | 0.018<br>(0.023) | -0.007<br>(0.022) | -0.016<br>(0.017) | 0.004<br>(0.020) | 6.836 | 0.001 | 0.268 | p = 0.009<br>d = 1.076 | p < 0.001<br>d = 1.676 | p = 0.367<br>d = 0.648 | p = 0.569<br>d = 0.445 | p = 0.603<br>d = -0.461 | p = 0.011<br>d = -1.033 |
| Medial-Lateral GRF PC1 | 79.04% | -0.190<br>(0.248) | -0.121<br>(0.217) | 0.071<br>(0.280) | -0.247<br>(0.328) | 3.732 | 0.016 | 0.167 | p = 0.888<br>d = -0.295 | p = 0.049<br>d = -0.948 | p = 0.959<br>d = 0.191 | p = 0.156<br>d = -0.752 | p = 0.626<br>d = 0.473 | p = 0.024<br>d = 1.026 |
| Rearfoot Eversion Angle PC2 | 8.04% | -3.051<br>(8.277) | -2.345<br>(7.191) | 6.473<br>(7.474) | -6.653<br>(6.941) | 7.537 | < 0.001 | 0.288 | p = 0.994<br>d = -0.090 | p = 0.008<br>d = -1.175 | p = 0.687<br>d = 0.446 | p = 0.006<br>d = -1.171 | p = 0.482<br>d = 0.586 | p < 0.001<br>d = 1.735 |

Additional variables were also unique to C3. There was a significant difference for the third PC of the knee abduction moment, where the PC score of C3 was significantly higher than C2 and C4, but not C1 (Supplementary Table 3, Supplementary Figure 3.A1), indicating less overall range over the stance phase (Supplementary Figure 3.A2). The score for C1 was also significantly higher than C2. The PC score of C3 was significantly higher than the other subgroups for the first PC of the vertical GRF (Supplementary Table 3, Supplementary Figure 3.B1), indicating greater overall magnitude during stance (Supplementary Figure 3.B2). Of note, the PC score for C2 was also higher than C4.

### Characterizing Cluster 4 (C4)

There was a significant difference for the first PC of the knee abduction angle, where the PC score of C4 was significantly higher than C2 and C3, but not C1 (Table 4, Figure 3.A1), indicating larger angle during stance (Figure 3.A2). There was also a significant difference for third PC of the hip adduction angle, where the PC score of C4 was significantly lower than other subgroups (Table 4, Figure 3.A1), indicating greater hip adduction in early stance and less range of motion (Figure 3.B2). Furthermore, there was a significant difference for the second PC of the hip rotation angle, where the PC score of C4 was significantly higher than C1, C2, and C3 (Table 4, Figure 3.C1), indicating greater internal rotation in early stance and greater external rotation in late stance (Figure 3.C2).

**Figure 3:**
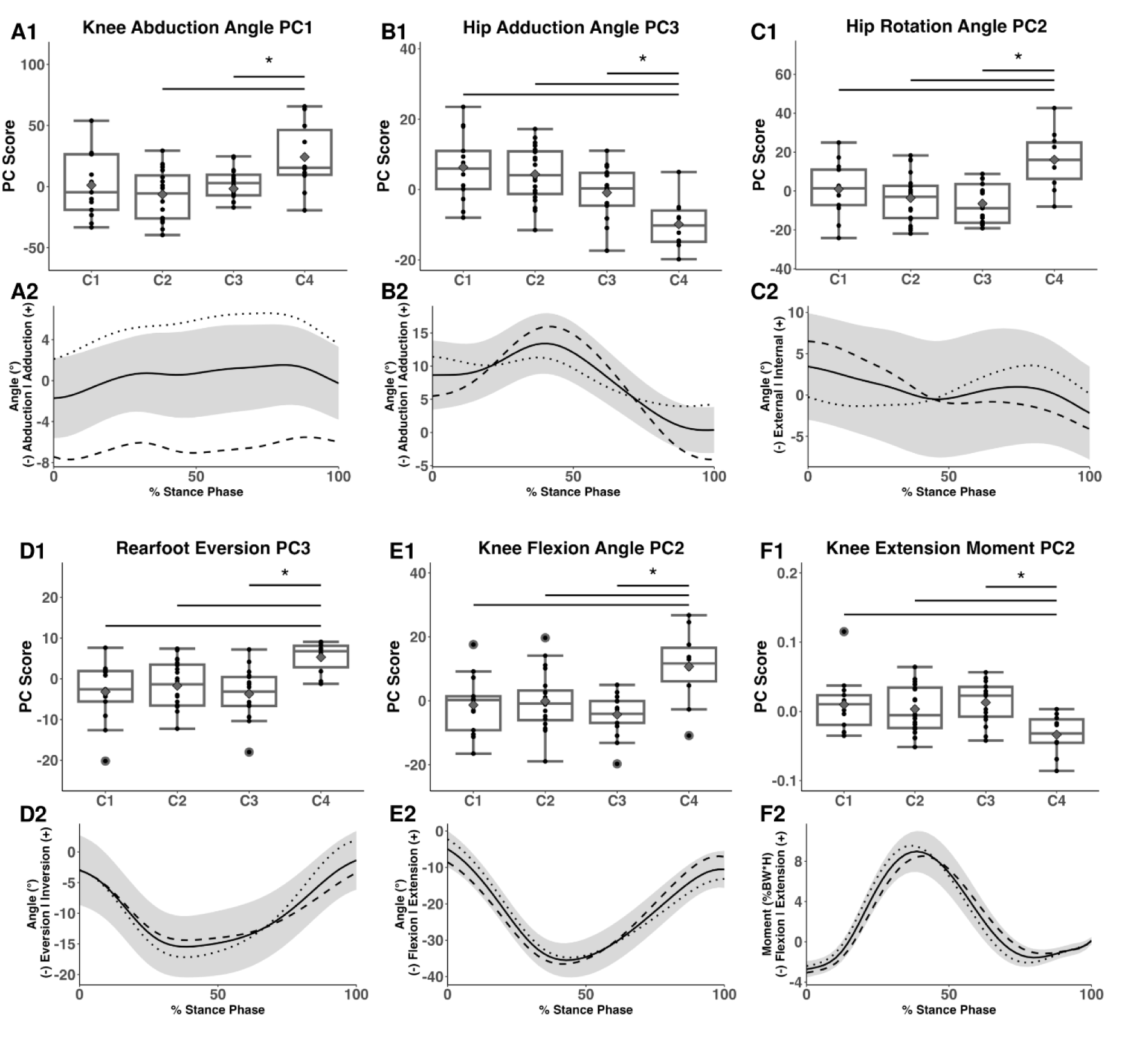
Box plots (top) and interpretation plots (bottom) for the kinematic and kinetic variables that characterized C4. Gray diamonds represent group means and black dots represent individual observations. * significant difference from other groups indicated by significance bars. In the interpretation plots, the dotted and dashed lines represent the discriminant vectors for the 5^th^ and 95^th^ percentile scores, respectively. PCs were interpreted by identifying deviations from the mean time-series signal shown in black with standard deviation represented by the gray shaded region.

**Table 4:** Explained variance, mean and standard deviation (SD) of PC scores, ANOVA results, and post-hoc testing results for the kinematic and kinetic variables that characterized C4.

|  |  | PC Score Mean (SD) |  |  |  | ANOVA Results |  |  | Post-Hoc Testing Results |  |  |  |  |  |
| --- | --- | --- | --- | --- | --- | --- | --- | --- | --- | --- | --- | --- | --- | --- |
| Variable | Explained Variance | C1 | C2 | C3 | C4 | F | p | $\eta^2$ | C1 vs. C2 | C1 vs. C3 | C1 vs. C4 | C2 vs. C3 | C2 vs. C4 | C3 vs. C4 |
| Knee Abduction Angle PC1 | 82.79% | 1.247<br>(25.421) | -6.328<br>(19.602) | -1.632<br>(19.305) | 24.279<br>(27.154) | 4.122 | 0.010 | 0.181 | p = 0.793<br>d = 0.333 | p = 0.987<br>d = 0.125 | p = 0.094<br>d = -0.840 | p = 0.926<br>d = -0.235 | p = 0.006<br>d = -1.320 | p = 0.033<br>d = -1.107 |
| Hip Adduction Angle PC3 | 4.18% | 6.323<br>(9.278) | 4.340<br>(7.642) | -0.871<br>(7.103) | -9.853<br>(6.771) | 9.473 | < 0.001 | 0.337 | p = 0.899<br>d = 0.231 | p = 0.082<br>d = 0.856 | p < 0.001<br>d = 1.866 | p = 0.212<br>d = 0.685 | p < 0.001<br>d = 1.862 | p = 0.034<br>d = 1.238 |
| Hip Rotation Angle PC2 | 9.78% | 0.971<br>(13.104) | -3.579<br>(12.058) | -6.493<br>(9.897) | 16.058<br>(14.096) | 7.564 | < 0.001 | 0.288 | p = 0.739<br>d = 0.353 | p = 0.378<br>d = 0.633 | p = 0.029<br>d = -1.064 | p = 0.895<br>d = 0.255 | p < 0.001<br>d = -1.485 | p < 0.001<br>d = -1.866 |
| Rearfoot Eversion Angle PC3 | 4.55% | -3.178<br>(7.160) | -1.694<br>(5.899) | -3.686<br>(5.927) | 5.311<br>(3.676) | 5.187 | 0.003 | 0.217 | p = 0.904<br>d = -0.224 | p = 0.996<br>d = 0.076 | p = 0.009<br>d = -1.374 | p = 0.757<br>d = 0.328 | p = 0.023<br>d = -1.286 | p < 0.003<br>d = -1.662 |
| Knee Flexion Angle PC2 | 11.42% | -1.327<br>(8.953) | -0.167<br>(8.875) | -4.32<br>(5.973) | 10.704<br>(10.929) | 6.268 | 0.001 | 0.251 | p = 0.983<br>d = -0.126 | p = 0.797<br>d = 0.390 | p = 0.011<br>d = -1.166 | p = 0.494<br>d = 0.526 | p = 0.013<br>d = -1.093 | p < 0.001<br>d = -1.770 |
| Knee Extension Moment PC2 | 14.31% | 0.01<br>(0.038) | 0.003<br>(0.033) | 0.013<br>(0.029) | -0.034<br>(0.028) | 4.622 | 0.006 | 0.198 | p = 0.949<br>d = 0.176 | p = 0.994<br>d = -0.092 | p = 0.016<br>d = 1.22 | p = 0.822<br>d = -0.298 | p = 0.030<br>d = 1.134 | p = 0.005<br>d = 1.582 |

C4 was also characterized by significantly higher scores for the third PC of the rearfoot eversion angle and second PC of the knee flexion angle compared to other subgroups (Table 4, Figures 3.D1 and 3.E1). These PCs indicated a smaller peak rearfoot eversion angle, less rearfoot range of motion in mid to late stance, and earlier peak knee flexion angles (Figures 3.D2 and 3.E2). Lastly, there was a significant difference for the second PC of the knee extension moment, where the PC score of C4 was significantly lower than C1, C2, and C3 (Table 4, Figure 3.F1), indicating an earlier peak extension moment (Figure 3.F2).

## Discussion

PFP is one of the most common running related injuries experienced by physically active individuals. While there is agreement that biomechanical factors play a central role in the development of PFP, there is also a lack of consensus for a single pathway to the injury. Rather, initial evidence suggests the existence of subgroups with unique PFP pathways. The aim of this study was to identify subgroups of individuals experiencing PFP during running, an activity known to contribute to pain development, and it was hypothesized that two to four subgroups with unique biomechanical patterns related to PFP development would exist. In support of this hypothesis, four subgroups were identified, three of which were dominated by symptomatic individuals who displayed unique movement patterns coinciding with pathways leading to development of PFP previously outlined in a pathomechanical model (14). Specifically, C1 displayed larger knee abduction moments and distal loading adaptations, C3 was characterized by distal kinematic adaptations and more medially directed ground reaction forces, and C4 was characterized by proximal kinematic adaptations. This suggests that findings of prior cross-sectional studies which found significant differences in biomechanics between healthy controls and those with PFP may have drawn from samples of injured individuals who belong to a specific subgroup. The robust approach taken in this study, and relevance of subgroups to the pathomechanical model (14), showcase potential for more targeted analysis in future work that may improve consistency of findings and enhance clinical applicability.

The first subgroup, C1, was characterized by greater magnitudes of knee abduction and rotation moments, and lower magnitude of the rearfoot inversion moment over the stance phase. Larger knee abduction impulses have been found in those with PFP compared to healthy controls, and knee abduction impulses predicted development of PFP in runners (20). However, another study found no differences in peak knee abduction moment between PFP and healthy controls during running, suggesting abnormally large frontal plane loading patterns may be a subgroup specific injury mechanism (16). Smaller rearfoot inversion moments were also found within this group, suggesting a potential distal loading factor contributing to local kinetic adaptations. Prior work has identified a connection between rearfoot inversion and knee abduction moments (46–49). Inverted, or medial, orthoses that were originally developed as a treatment option for runners who did not respond to standard orthotic treatment (48) significantly increased the peak knee abduction moment and reduced rearfoot inversion moments (50). Lateral and medial wedges were also associated with larger and smaller rearfoot inversion moments, respectively, and the peak knee abduction moment decreased as wedges progressed from medial to lateral (49). While orthotic or footwear modification have been explored as a treatment for PFP, there is lack of strong evidence to support their efficacy as a long-term solution (51). The identified cluster with unique characteristics related to the knee abduction impulse and rearfoot inversion moments may be important to consider for future studies and the development of footwear treatment approaches.

The third subgroup, C3, was characterized by an earlier peak timing and greater loading rate of the knee abduction moment, more medially directed GRFs, and greater range of rearfoot motion in the frontal plane. Sagittal plane loading rates have been reported to be higher in those with PFP compared to healthy controls (52) but limited data are available for the frontal plane loading rates. Peak knee abduction moments were greater, and stance time was shorter, in females who experienced traumatic joint injury, which may indicate that larger frontal plane loading rates are a risk factor for knee injury (53). Therefore, greater loading rates in the frontal plane may be related to PFP development in this subgroup, yet this factor is not well understood but it may stem from distal kinematic adaptations. Rodrigues et al. (2013) found that runners experiencing anterior knee pain used a significantly higher percentage of available rearfoot eversion range of motion compared to healthy controls. Greater rearfoot eversion angle is associated with tibial internal rotation and can disrupt the screw home mechanism, contributing to knee valgus and increased knee abduction moments (20,54–56). Excessive subtalar pronation coupled with internal tibial rotation is one of the most widely recognized distal PFP pathways, however prior work has not found this as a consistent biomechanical factor in this population (14,57). This subgroup also demonstrated a greater magnitude of rearfoot eversion angular velocity in early stance which aligns temporally with the earlier peak knee abduction moment, suggesting a possible relationship between distal kinematic and local kinetic frontal plane patterns. In addition, more medially directed GRFs over the stance phase can result in a larger lever arm between the knee joint center and the line of action of the GRF that can contribute to a greater magnitude of the peak knee abduction moment (20). Overall, the distal kinematic pattern and GRF direction unique to this group may have impacted frontal plane knee joint loading patterns that could contribute to elevated patellofemoral joint stress. Notably, there were five healthy individuals in this subgroup, and it is of interest to determine if they may be at risk of developing PFP if these biomechanical factors are not addressed.

The fourth subgroup, C4, was primarily characterized by distinct proximal kinematics including greater hip internal rotation and adduction in early stance and less frontal plane hip range of motion throughout the stance phase. However, C4 also displayed larger knee abduction angles, lower peak rearfoot eversion angles, less overall frontal plane foot range of motion, and earlier timing of the peak knee flexion angle and knee extension moment. Prior work has reported correlations between frontal and transverse hip kinematics, resulting in larger knee abduction angles (58). Furthermore, larger knee abduction angles have been associated with diminished patellofemoral joint contact area and increased patellofemoral joint stress (59,60). Proximal adaptations such as greater hip internal rotation and adduction have been found in those with PFP compared to healthy controls (61,62), however findings are not consistent across all studies (14,21). Therefore, this proximal pathway is likely subgroup specific, as demonstrated by this data-driven clustering approach. Notably, this group contained predominantly females, which is not surprising given that females tend to exhibit greater hip adduction than males during running (63). However, not all females were assigned to this subgroup, suggesting that proximal kinematic adaptations are not the only female specific PFP pathway. The lower peak rearfoot eversion angle and less frontal plane range of motion in this subgroup further supports a unique proximal kinematic pathway. Interestingly, this was the only subgroup that demonstrated sagittal plane adaptations, which may coincide with findings of a recent meta-analysis (21). The earlier peak timing of both the knee flexion angle and knee extension moment is consistent with prior findings of larger sagittal plane loading rate in those with PFP (52).

Optimizing the clusters with CPI and the feature selection process was key to uncovering these subgroups. By maximizing CPI across several feature sets, a cluster with as many healthy individuals and as few symptomatic individuals as possible was generated. Gini impurity was provided as a well-established measure of cluster purity, however optimization with this metric may have resulted in a single cluster with all individuals with PFP and no healthy controls. In this scenario, biomechanical features unique to PFP subgroups may not have been revealed. However, the moderate level of purity (CPI: 0.475, Gini: 0.330) and moderate degree of separation and compactness (Dunn: 0.413) of the clusters suggest some overlap between the healthy and symptomatic groups. While the clusters captured meaningful distinctions, the presence of cross-over indicated that some individuals may have exhibited biomechanical patterns of multiple subgroups. The characterization process was particularly important as it highlights the unique attributes of each subgroup beyond the scope of a clustering model tasked with providing label assignment. These findings underscore the complex etiology of PFP and the need for further refinement in future approaches.

The lack of differences for variables in the REPORT-PFP Checklist (29) suggests that biomechanical adaptations should be given consideration when designing and implementing targeted treatments. This brings into question how subgrouping could be performed in a clinical setting without motion capture equipment and may require something more translational such as wearable sensors and/or marker less motion capture.

This study did not come without limitations. First off, we did not account for other factors associated with development of PFP such as muscle function, soft tissue restraints, and altered tibiofemoral joint anatomy. Measures of muscular strength, electromyography, and imaging modalities would be required to quantify these PFP related factors. Variables representing such factors may have facilitated more clear and concise subgroups. Furthermore, some subgroups, specifically C2 and C3 resulted in higher Gini impurity, suggesting clusters were not optimally pure. Ideally, this metric would be closer to 0 indicating clusters with less mixture of healthy and symptomatic individuals. Additionally, patellofemoral joint loads were not estimated, which may have added robustness to biomechanical and clustering analyses. Lastly, the number of clusters was maximized at four. It is possible that more clusters exist, however larger sample sizes would be required to provide enough statistical power to detect differences between subgroups.

In conclusion, this data-driven approach leveraging PCA and a clustering approach optimized on an index of cluster purity, which implicitly accounted for injury status, resulted in four distinct clusters. Three of these clusters contained mostly symptomatic individuals, each with unique biomechanical factors associated with PFP. These subgroups may benefit from targeted treatment approaches that have shown to be more effective compared to standardized interventions (2), especially if they are identifiable in clinical settings with the use of other modalities such as wearable sensors and supervised machine learning models.

## Acknowledgements

This study was funded by the University of Massachusetts Priscilla M. Clarkson Graduate Scholarship in Kinesiology.

## Data Availability Statement

The datasets generated during and/or analyzed during the current study are not publicly available because the datasets have not been deposited into a public repository but are available from the corresponding author by emailing the corresponding author, Katherine A. Boyer.

